# The impact of thermal pasteurization on viral load in human milk and other matrices: A rapid review

**DOI:** 10.1101/2020.05.23.20111369

**Authors:** Michael A. Pitino, Deborah L. O’Connor, Allison J. McGeer, Sharon Unger

**Affiliations:** Department of Nutritional Sciences, University of Toronto, Toronto, Canada; Translational Medicine Program, The Hospital for Sick Children; Department of Microbiology, Sinai Health, Toronto, Canada; Department of Laboratory Medicine and Pathobiology, University of Toronto, Toronto, Canada; Dalla Lana School of Public Health, University of Toronto, Toronto, Canada; Division of Neonatology, The Hospital for Sick Children, Toronto, Canada; Department of Pediatrics, Sinai Health, Toronto, Canada; Department of Pediatrics, University of Toronto, Toronto, Canada

**Keywords:** viral infectivity, viruses, Holder pasteurization, thermal pasteurization, human milk, donor milk, milk banking, SARS-CoV-2

## Abstract

Holder pasteurization (62.5°C, 30 min) of human milk (HM) is thought to reduce the risk of transmitting viruses to an infant. Some viruses may be secreted into milk – others may be contaminants. The effect of thermal pasteurization on viruses in HM has yet to be rigorously reviewed. The objective of this study is to characterize the effect of commonly used pasteurization techniques on viruses in HM and non-HM matrices. Databases (MEDLINE, Embase, Web of Science) were searched from inception to April 20^th^, 2020 for primary research articles assessing the impact of pasteurization on viral load or detection of live virus. Reviews were excluded, as were studies lacking quantitative measurements or those assessing pasteurization as a component of a larger process. Overall, 65,131 reports were identified, and 108 studies included. Pasteurization of HM at a minimum temperature of 56°C-60°C is effective at reducing detectable live virus. In cell culture media or plasma, coronaviruses (e.g., SARS-CoV, SARS-CoV-2, MERS) are highly susceptible to heating at ≥56°C. Although pasteurization parameters and matrices reported vary, all viruses studied, with the exception of parvoviruses, were susceptible to thermal killing. Future research important for the study of novel viruses should standardize pasteurization protocols and should test viral inactivation using a human milk matrix.

Novelty bullets

- In all matrices, including human milk, pasteurization at temperatures of 62.5°C was generally sufficient to reduce surviving viral load by several logs or to below the limit of detection.
- Holder pasteurization (62.5°C, 30 min) of human donor milk should be sufficient to inactivate non-heat resistant viruses, including coronaviruses, if present.

## Introduction

Since the emergence of SARS-CoV-2 in late 2019, ensuring that current high-quality screening, handling and pasteurization standards are sufficient for maintaining a safe supply of human donor milk has been an ongoing challenge for milk banks(Furlow 2020). Human donor milk is used as a bridge for hospitalized infants while their mother’s own milk supply is being established; among very low birth weight infants, the use of human donor milk instead of preterm formula as a bridge has been shown to reduce the incidence of necrotizing enterocolitis(Underwood 2013; Quigley et al. 2019). Milk banking associations, including the Human Milk Banking Association of North America and the European Milk Banking Association have responded to the pandemic by issuing new guidelines with respect to enhanced donor screening, including asking specific questions to assess the likelihood of a potential donor being infected with SARS-CoV-2(“COVID-19: EMBA Position Statement” 2020; “Milk Banking and COVID-19” 2020). While all donor milk from non-profit milk banks in North America is pasteurized using the Holder method (62.5°C, 30 min) to inactivate potentially pathogenic bacteria and viruses, additional research is warranted to determine whether SARS-CoV-2, is inactivated by Holder pasteurization(Arslanoglu et al. 2010; *Guidelines for the Establishment and Operation of a Donor Human Milk Bank* 2018).

At present, the virome of human milk has been understudied. Few studies have investigated whether or not viruses that may cause disease in preterm infants are present in human milk(Mohandas and Pannaraj 2020). Viruses may be present in human milk as a result of secretion into the milk in the mammary tissue, notably, cytomegalovirus, human t-lymphocytic virus, and human immunodeficiency virus (HIV), or may be present as a contaminant from skin or respiratory droplets either in the milk or on the containers(Michie 2001). Regardless of origin, it is important to understand how viruses found in human milk respond to thermal pasteurization. To date, there has been no systematic review of the impact of thermal pasteurization on viral load or live virus detection in a human milk matrix or other non-human milk matrices. The primary aim of this review is to characterize studies conducted in human milk to determine how certain viral families that are either present in human milk, or used as surrogates, respond to thermal pasteurization as assessed by viral load or live virus detection. To expand the scope of viruses tested, the secondary objective is to summarize studies conducted in non-human milk matrices that have examined the effect of thermal pasteurization on any virus. This review also aims to compare viruses that have been assessed in studies using both human milk and non-human milk matrices to ascertain any trends in susceptibility to thermal pasteurization.

## Materials and methods

The Preferred Reporting Items for Systematic Reviews and Meta-Analyses (PRISMA) guidelines were followed in completion of this rapid review, except where indicated (Moher et al. 2009). This rapid review is in response to the COVID-19 pandemic.

### Search strategy and selection criteria

References for this rapid review were identified through electronic searches of various online databases including MEDLINE, Embase and Web of Science, from database inception to April 20^th^, 2020, with the assistance of a research librarian. The search strategy focused on keywords to identify articles that assessed the effect of thermal pasteurization or heat inactivation, including Holder pasteurization, on the detection of live virus or viral load in human milk or other non-human milk matrices. The names of viral families, as per the current taxonomic classification, were included in the search as they may be present in human milk (by secretion or contamination) or could be used as surrogate viruses to model highly pathogenic or non-culturable viruses(King et al. 2012).

The keywords and MeSH terms included for all database searches were intended to capture all relevant research with respect to thermal pasteurization of viruses in human milk, the primary outcome of this rapid review. To increase the scope, we supplemented the search to capture research articles that tested all matrices other than human milk. The search strategy is summarized in Table S1 and included three main ideas. The first concept included viral taxonomic families using keywords and MeSH terms based on the nomenclature suggested by the International Committee on Taxonomy(King et al. 2012). The second concept consisted of synonyms and phrases closely related to human milk (e.g. breast milk, donor milk etc.). Lastly, the final concept was thermal pasteurization and its synonyms (e.g., Holder pasteurization, heat etc.). Our initial search aimed to retrieve articles specific to human milk which was achieved by combining all three concepts; by only retaining the first and last concept, a second set of articles was retrieved that theoretically involved thermal pasteurization and viruses in all other matrices, including human milk. Grey literature was searched as per the previously published guidelines including from dissertations and google advanced search(Natal 2019). Articles resulting from those searches and relevant references cited in those articles were reviewed.

After duplicates were removed, titles, then abstracts were screened by a single reviewer. Primary research articles were included if they assessed the effect of Holder pasteurization (62.5°C −63°C) or any other heat treatment on viral load or detection of live virus in human milk or other matrices. Eligible study designs included pre-post or longitudinal; in either design, the outcome, detection of live virus or viral load, was assessed before and after pasteurization, or at discrete time points during a given pasteurization process. Qualitative, observational and review studies were excluded, in addition to experimental studies that did not assess viral load (quantitative) or detectable live virus. Studies that investigated how thermal pasteurization and the addition of matrix stabilizers, affects viral load or live virus detection were also excluded; the outcome of these studies may be confounded by the fact that the integrity of viruses may be different as certain stabilizers are added or removed. Studies were also excluded if thermal pasteurization was tested in combination with other processing techniques, (e.g., irradiation, lyophilization during the production of plasma concentrates), unless the study was appropriately controlled. The primary rationale being that aspects of processing, other than heat, may also lead to the inactivation of viruses. Reports on clinical trials or studies published in non-scientific journals were not included. All studies irrespective of language or year published were included.

Multiple attempts were made to retrieve the full-text of all articles screened on the basis of title and abstract including interlibrary loan and/or author follow-up. Data were extracted from eligible full-text articles including viruses tested, matrix used, thermal pasteurization parameters (temperature, time) and a measure of reduction in viral load/detectable virus. Included studies were summarized after being dichotomized into two groups depending on whether detectable live virus or viral load was tested in human milk or another matrix. To determine whether a human milk matrix affected the results, a subanalysis was conducted on studies that tested the same viruses in both human milk and non-human milk matrices. In this subanalysis, only studies that assessed virus presences by plaque reduction assay or endpoint dilution (TCID_50_) were included. First, viruses that were tested in both groups were determined by cross-referencing; relevant data (log-reduction, temperature, and duration of pasteurization) was then extracted and aggregated. Unless otherwise defined, complete inactivation is a concentration of virus that was below the lower detection limit of the assay. If multiple studies assessed the same virus, the pasteurization conditions used in the summary were matched as closely as possible to the data available in studies experimenting with human milk.

## Results

### Study selection and characteristics

The selection of studies is summarized in Figure 1. A total of *n=*65,131 reports were identified and assessed for eligibility. This included 23,441 citations from MEDLINE, 34,479 citations from Embase, 7,200 records from Web of Science and 11 from manual searches. Altogether, *n*=64,950 records were excluded on the basis of title and abstracts alone, encompassing articles that did not meet the inclusion criteria (*n=*44,287 records) or duplicate records (*n=*20,663). After title and abstract screening, 181 reports remained for full-text review. Upon full-text review, 73 reports were excluded: 6 were duplicate records, *2* could not be retrieved and 65 did not meet inclusion criteria. Thus, 108 articles were included in the review and were organized according to the matrix used in testing the effect of pasteurization on viral load.

**Figure 1.**
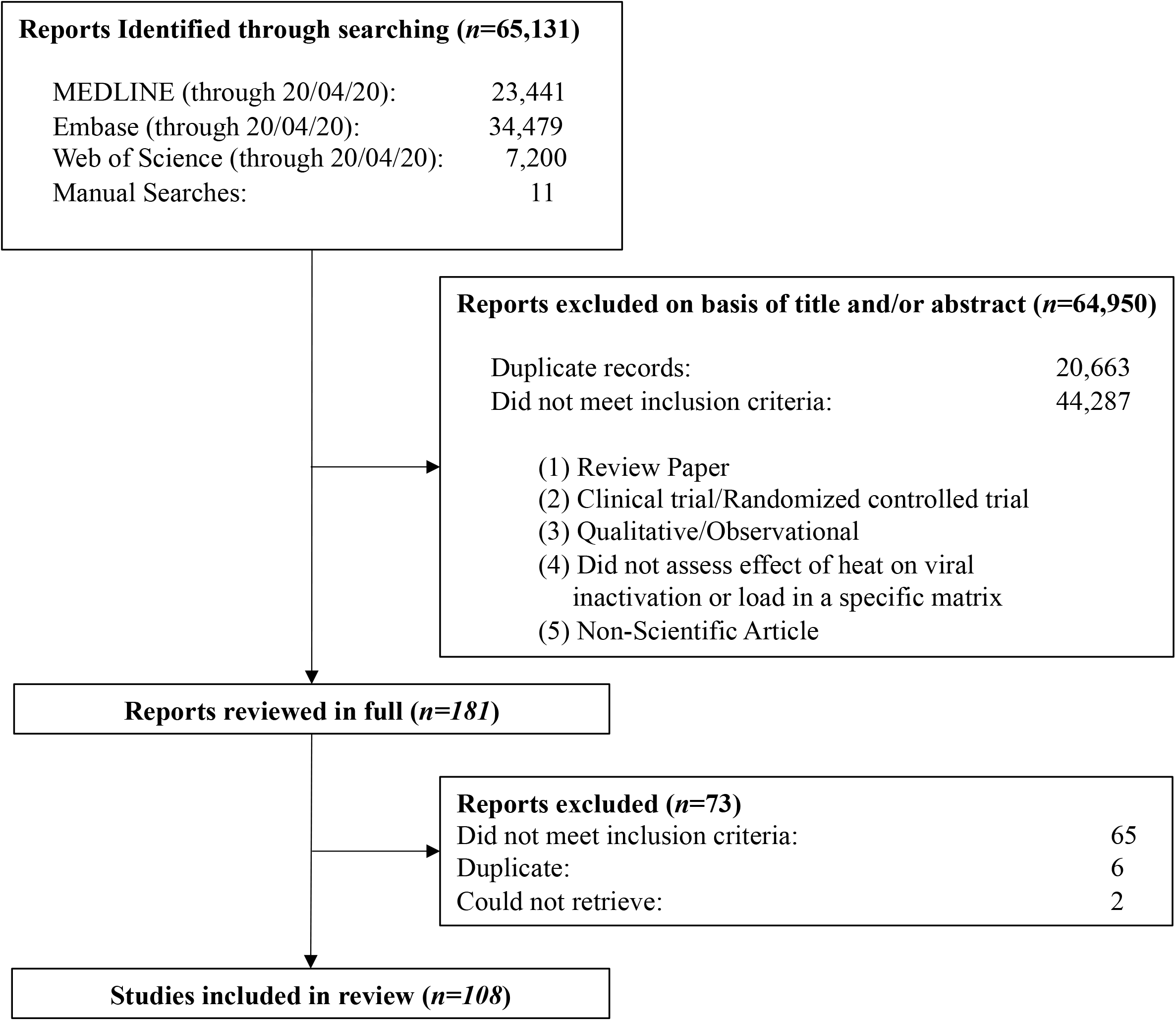
PRISMA flow diagram describing the selection of studies for inclusion in the review

### Studies conducted in human milk

First, we summarized 18 unique studies that used human milk as the matrix to test the effect of pasteurization on thirteen different viruses (Table 1). Most studies reported on viral addition experiments, while few studies subjected milk with endogenous virus to thermal pasteurization. Predominantly, the viruses tested were caspid enveloped and belonged to 8 different viral families including: caliciviridae, filoviridae, flaviviridae, herpesviridae, papillomaviridae, picornaviridae, retroviridae, and togaviridae. Cytomegalovirus and HIV were the most common viruses studied with 8 and 7 articles respectively. To assess surviving virus concentration following pasteurization, plaque reduction assays and endpoint titration assays (TCID_50_) were most frequently used, although some studies used immunofluorescence, reverse-transcriptase enzymatic assays and secreted embryonic alkaline phosphatase (SEAP) reporter assay.

**Table 1.** Summary of studies assessing the effect of heat, including Holder pasteurization, on viral inactivation in human milk

| Family | Virus | Envelope | Pasteurization | Method | Result | Reference |
| --- | --- | --- | --- | --- | --- | --- |
| Caliciviridae | Feline Calicivirus | No | 35°C-70°C, 2 min | PRA | >4.5-log PFU/mL reduction at 60°C, 2min; >5-log PFU/mL reduction (complete inactivation) at 65°C, 2 min | (Topping et al. 2009) |
| Filoviridae | Ebola virus | Yes | 62.5°C, 30 min | PRA | >5-log PFU/mL reduction (complete inactivation) at 62.5°C, 30 min | (Hamilton Spence et al. 2017) |
|  | Marburg virus | Yes | 62.5°C, 30 min | PRA | >5-log PFU/mL reduction (complete inactivation) at 62.5°C, 30 min | (Hamilton Spence et al. 2017) |
| Flaviviridae | Bovine viral diarrhea virus | No | 72°C, 16 sec | TCID<sub>50</sub> | >7-log TCID<sub>50</sub>/mL (complete inactivation) at 72°C, 4 sec | (Terpstra et al. 2007) |
|  | Zika virus | Yes | 63°C, 30 min | TCID<sub>50</sub> | >6-log TCID<sub>50</sub>/mL reduction (complete inactivation) at 63°C, 30 min | (Pfaender et al. 2017) |
| Herpesviridae | Cytomegalovirus | Yes | 55°C-72°C, 5 sec | TCID<sub>50</sub> | >4-log TCID<sub>50</sub>/mL reduction (complete inactivation) at 60°C, 5s | (Maschmann et al. 2019) |
| Herpesviridae | Cytomegalovirus | Yes | (1) 62°C, 2-15 sec; (2) 72°C, 5-15 sec; (3) 62°C, 30 min | Early antigen IF | Complete inactivation (no IEA + cells) for all treatments | (Klotz et al. 2018) |
| Herpesviridae | Cytomegalovirus | Yes | 62.5°C, 30 min | SEAP Reporter | Complete inactivation at 62.5°C, 30 min | (Donalisio et al. 2014) |
| Herpesviridae | Cytomegalovirus | Yes | (1) 62.5°C, 30 min; (2) 72°C, 5 sec | Early Antigen IF | Complete inactivation (no IEA+ cells) at 62.5°C, 30 min or 72°C, 5 sec | (Hamprecht et al. 2004) |
| Herpesviridae | Cytomegalovirus | Yes | (72°C, 87°C), 1-15 sec | PRA | >5-log PFU/mL reduction (complete inactivation) at 72°C, 5 sec or 87°C, 5 sec | (Goldblum et al. 1984) |
| Herpesviridae | Cytomegalovirus | Yes | 56°C, 62°C, 30 min | Cell culture toxicity | Complete inactivation (no cell culture toxicity) at 62°C, 30 min | (Dworsky et al. 1982) |
| Herpesviridae | Cytomegalovirus | Yes | 63°C, 1-16 min | PRA | 3.6-log PFU/mL reduction (complete inactivation) at 63°C, 8 min | (Friis and Andersen 1982) |
| Herpesviridae | Cytomegalovirus | Yes | (1) 56°C, 30 min; (2) 63°C, 30 min; (3) 100°C, 5 min | Cell culture toxicity | Complete inactivation (no detectable cytopathic effect) at 63°C, 30 min | (Welsh et al. 1979) |
| Herpesviridae | Herpes simplex virus | Yes | (1) 56°C, 30 min; (2) 63°C, 30 min; (3) 100°C, 5 min | PRA | 4.2-log PFU/mL reduction at 63°C, 30min; >7-log PFU/mL reduction (complete inactivation) at 100°C, 5 min | (Welsh et al. 1979) |
| Herpesviridae | Pseudorabies virus | Yes | 72°C, 16 sec | TCID<sub>50</sub> | >8-log TCID<sub>50</sub>/mL reduction (complete inactivation) at 72°C, 4 sec | (Terpstra et al. 2007) |
| Papillomaviridae | Human papillomavirus | No | 62.5°C, 30 min | SEAP Reporter | Complete inactivation at 62.5°C, 30 min | (Donalisio et al. 2014) |
| Parvoviridae | Porcine parovirus | No | 72°C, 16 sec | TCID<sub>50</sub> | <1-log TCID<sub>50</sub>/mL reduction at 72°C, 16 sec | (Terpstra et al. 2007) |
| Picornaviridae | Coxsackievirus B4 | No | (1) 56°C, 30 min; (2) 63°C, 30 min; (3) 100°C, 5 min | PRA | 3.8-log PFU/mL reduction at 56°C, 30 min; 3.6-log PFU/mL reduction at 63°C, 30 min; >7-log PFU/mL reduction at 100°C, 5 min | (Welsh et al. 1979) |
| Picornaviridae | Hepatitis A virus | No | 72°C, 16 sec | TCID<sub>50</sub> | 3.5-log TCID<sub>50</sub>/mL reduction at 72°C, 16 sec | (Terpstra et al. 2007) |
| Retroviridae | HIV-1 | Yes | 54°C -57°C, 33 min | RT activity | 4-log reduction (complete inactivation) at 56°C, 33 min | (Eglin, RP, Wilkinson 1987) |
| Retroviridae | HIV-1 | Yes | 55°C-70°C, time to max temperature | GFP indicator cells | 4-log IU/mL reduction (complete inactivation, no GFP+ cells) at 65°C, 5 sec | (Hoque et al. 2013) |
| Retroviridae | HIV-1 | Yes | 55°C-70°C, time to max temperature | PBMC neutralization assay | Complete inactivation after flash-heat treatment | (Volk et al. 2010) |
| Retroviridae | HIV-1 | Yes | Flash heating (>56°C for 6 min (peak 73°C)) | RT activity | >3.4-log copies/mL reduction (complete inactivation) after flash heating | (Israel-Ballard et al. 2007) |
| Retroviridae | HIV-1 | Yes | 72°C, 16 sec | TCID<sub>50</sub> | >8-log TCID<sub>50</sub>/mL reduction (complete inactivation) at 72°C, 4 sec | (Terpstra et al. 2007) |
| Retroviridae | HIV-1 | Yes | 56°C- 62.5°C, 12-15 min | RNA assay | >5-log copies/mL reduction (complete inactivation) after pasteurization | (Jeffery et al. 2001) |
| Retroviridae | HIV-1 | Yes | 62.5°C, 30min | TCID<sub>50</sub> | >5.5-log TCID<sub>50</sub>/mL reduction (complete inactivation) at 62.5°C, 30 min | (Orloff et al. 1993) |
| Togaviridae | Semliki forest virus | Yes | (1) 56°C, 30 min; (2) 63°C, 30 min; (3) 100°C, 5 min | PRA | 4.2-log PFU/mL reduction at 63°C, 30 min; >7-log PFU/mL reduction (complete inactivation) at 100°C, 5 min | (Welsh et al. 1979) |

**Note.** Human immunodeficiency virus, HIV; Immunofluorescence, IF; Green fluorescence protein, GFP; plaque forming unit, PFU; Peripheral blood mononuclear cell, PBMC; Plaque reduction assay, PRA; Reverse transcriptase, RT; Secreted embryonic alkaline phosphatase, SEAP; Tissue culture infectious dose 50, TCID<sub>50</sub>. Complete inactivation refers to a viral load that is below the detectable limit of the assay, unless otherwise noted.

Based on the literature reviewed, Holder pasteurization, defined as a temperature of 62.5°C −63°C held for 30 minutes, resulted in complete inactivation of viruses in the herpesviridae family, including cytomegalovirus (Dworsky et al. 1982; Hamprecht et al. 2004; Donalisio et al. 2014); however, complete inactivation of herpes simplex virus did not occur, requiring a temperature of 100°C, 5 min (Welsh et al. 1979). In fact, for cytomegalovirus specifically, some studies reported complete inactivation at 60°C-63°C for 5 seconds to 30 minutes(Friis and Andersen 1982; Klotz et al. 2018; Maschmann et al. 2019). Similarly, retroviridae were susceptible to heating in a human milk matrix whereby complete inactivation was observed after pasteurization above 60°C, for a minimum of 5 seconds. In particular, flash heating and Holder pasteurization completely inactivated HIV-1 in human milk(Orloff et al. 1993; Israel-Ballard et al. 2007; Volk et al. 2010; Hoque et al. 2013); high temperature short time (72°C for 8 seconds) similarly yielded complete inactivation (>5.5-log reduction)(Terpstra et al. 2007). Holder pasteurization was found to inactivate (>5-log reduction) Ebola virus and Marburg virus of the filoviridae family, Zika virus (>6-log reduction) of the flaviviridae family, Semliki forest virus of the togaviridae family (4.2-log reduction) and human papillomavirus of the papillomaviridae family(Welsh et al. 1979; Hamilton Spence et al. 2017; Pfaender et al. 2017). Some non-enveloped members of the picornaviridae family were found to be more resistant to heating(Terpstra et al. 2007); flash heating (72 °C for 16 seconds) of hepatitis A virus and porcine parovirus yielded a 2-or 0.5-log reduction in TCID_50_/mL respectively. Infectivity of coxsackievirus persisted after Holder pasteurization, although reduced by 3.6-log PFU/mL(Welsh et al. 1979).

### Studies conducted in non-human milk matrices

Second, we summarized the remaining 90 unique studies that were identified during the literature review that assessed the effect of thermal pasteurization on viruses in a non-human milk matrix (Table 2). Cell culture media was the most prevalent matrix used in testing; other common matrices included bovine milk, bovine serum, human serum albumin, human plasma. In total, 21 unique families of viruses were tested including: adenoviridae, anelloviridae, birnaviridae, caliciviridae, circonviridae, coronaviridae, flaviviridae, hepadnaviridae, hepeviridae, herpesviridae, orthomyxoviridae, paramyxoviridae, parvoviridae, picornaviridae, polymaviridae, poxviridae, reoviridae, retroviridae, rhabdoviridae, togaviridae. The majority of studies tested non-enveloped viruses in the families of picornaviridae (*n=38*), and caliciviridae (*n=24*), in addition to retroviridae (*n=16*).

**Table 2.**
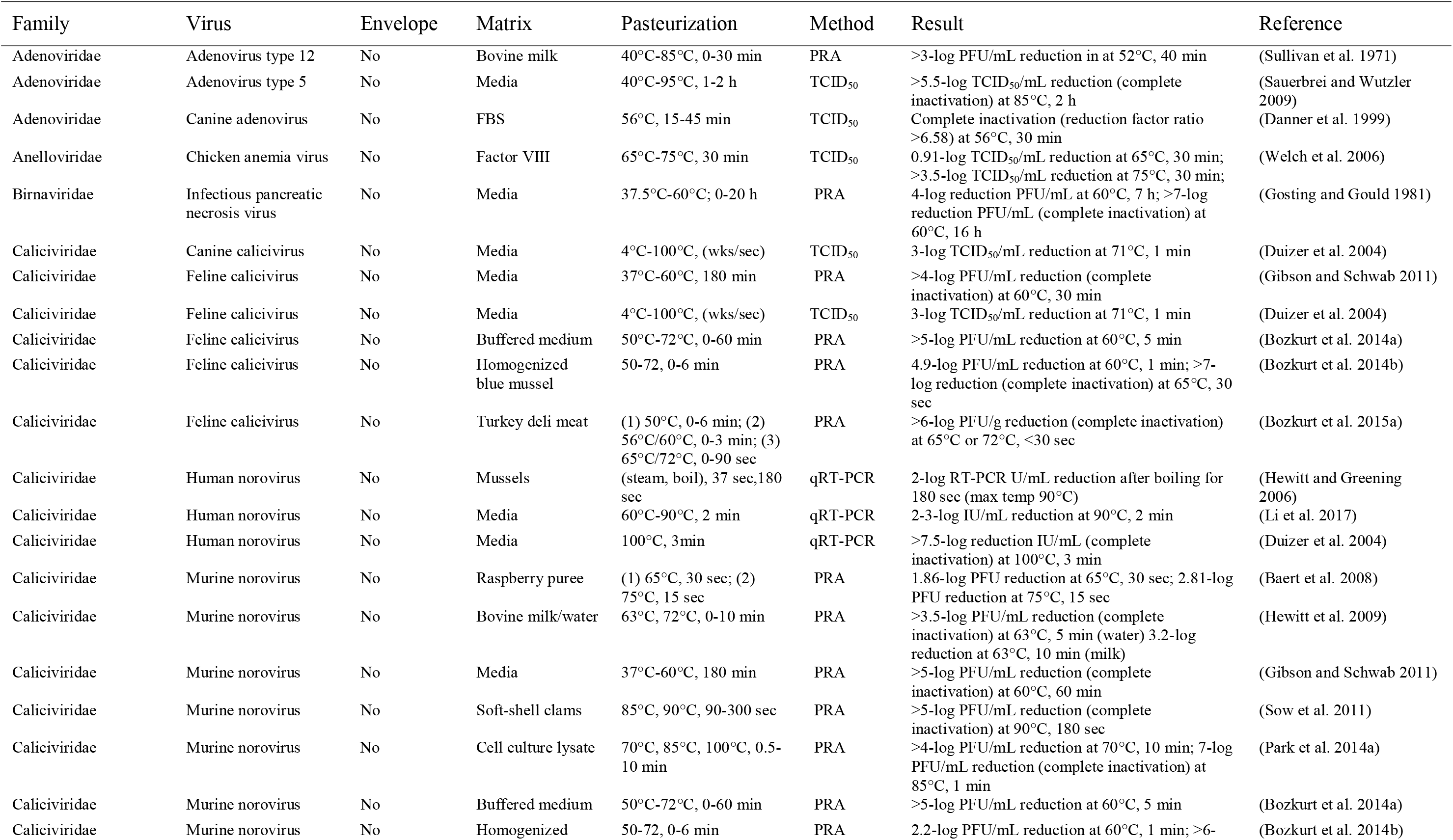
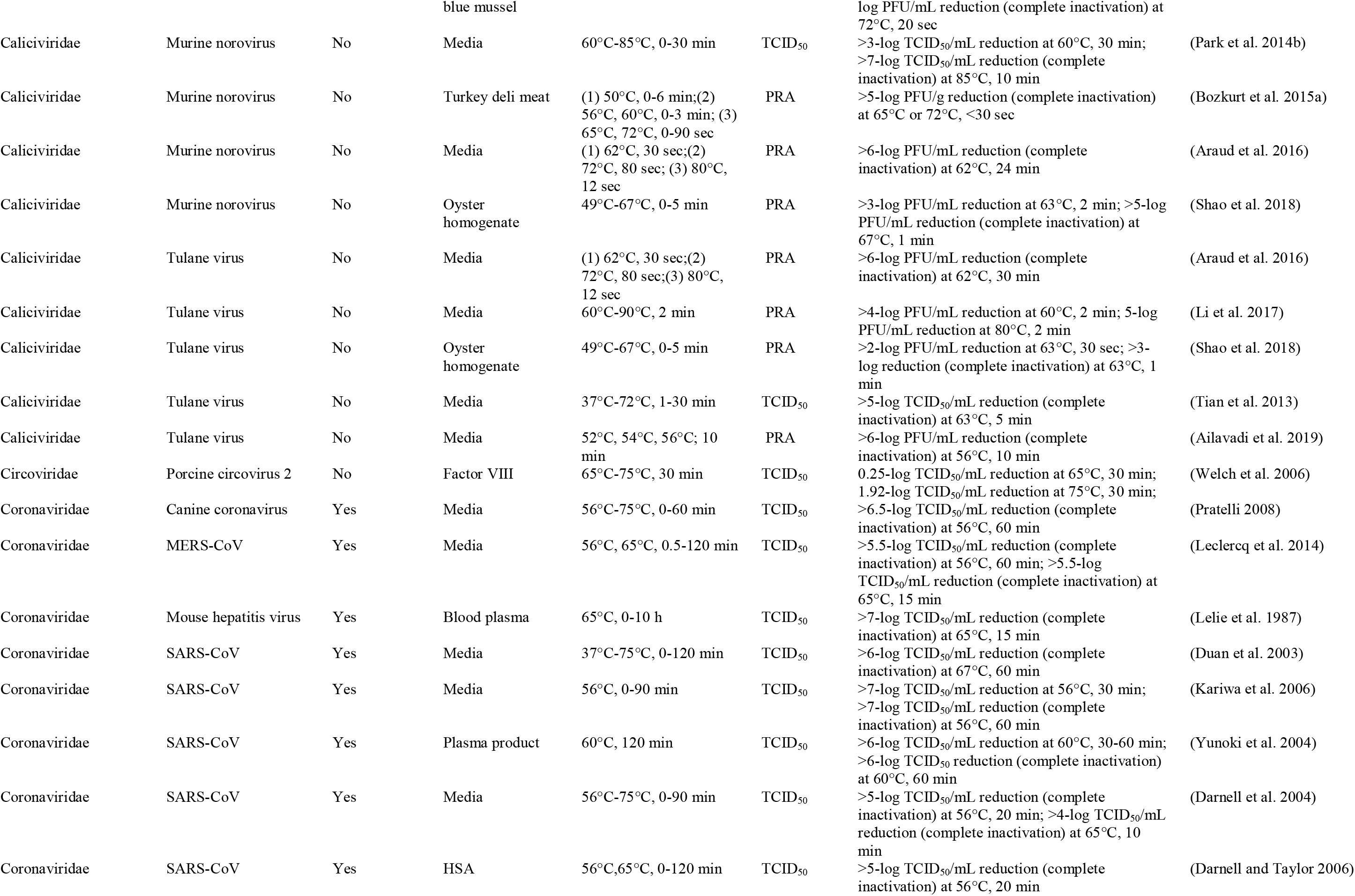
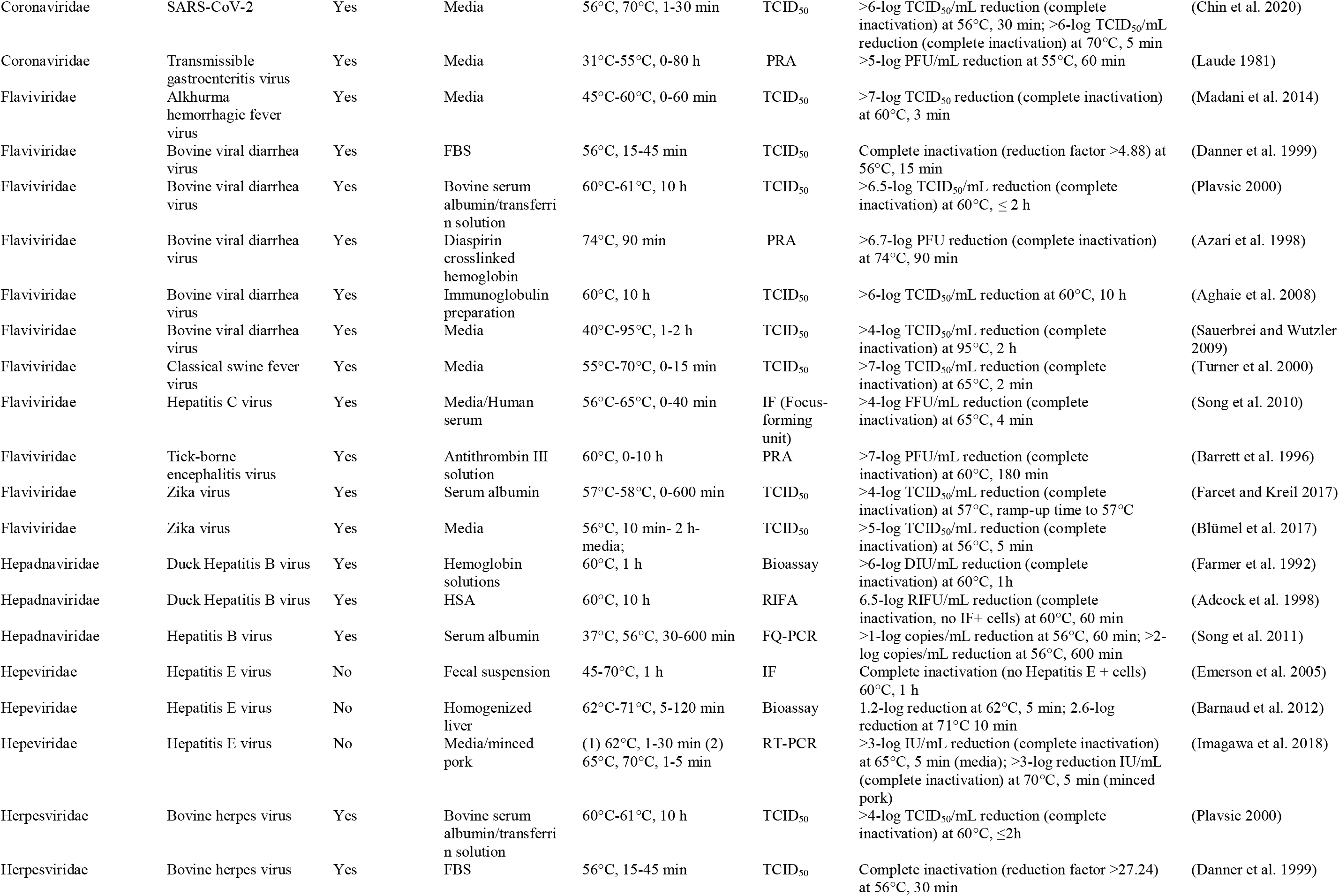
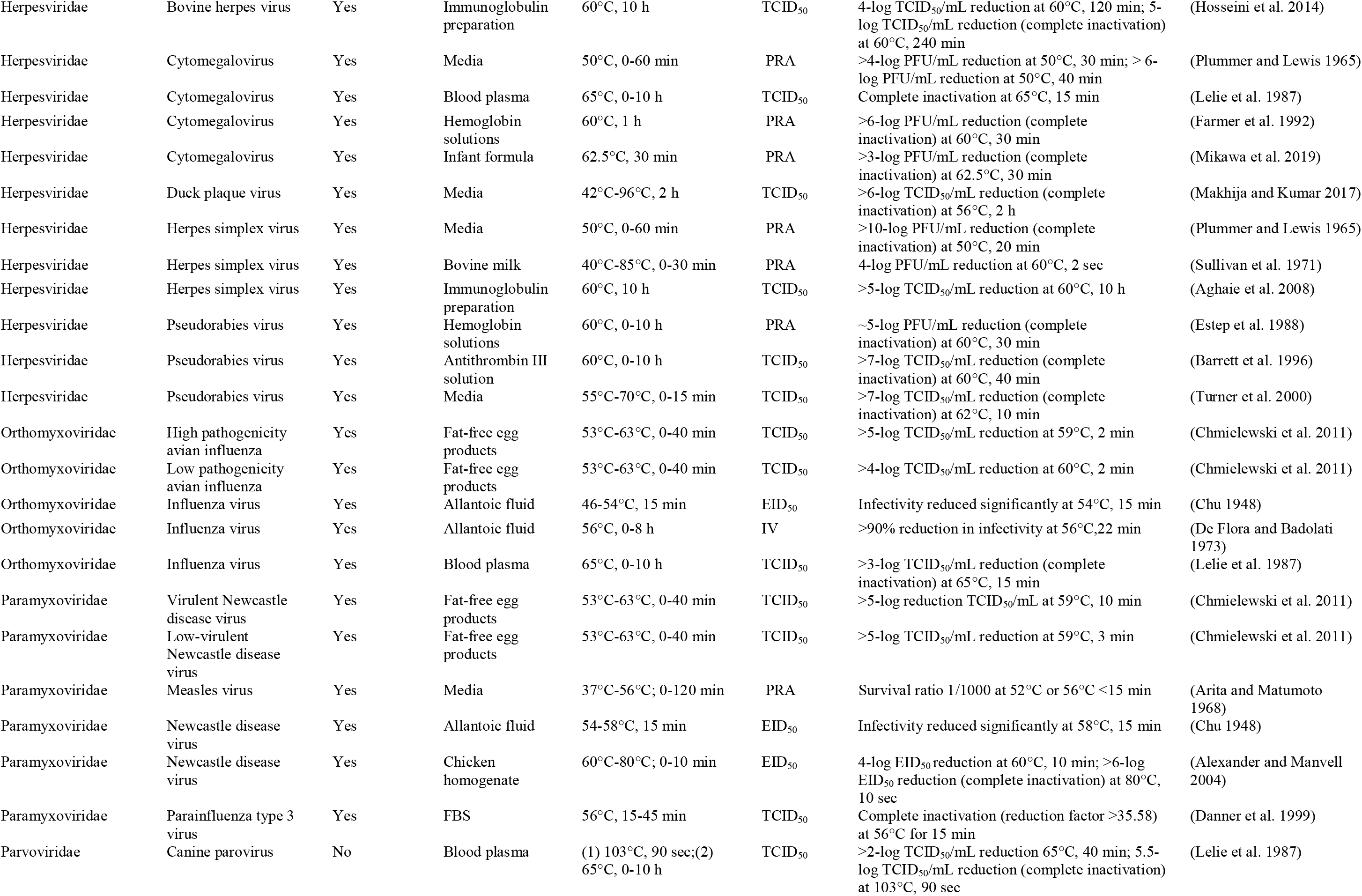
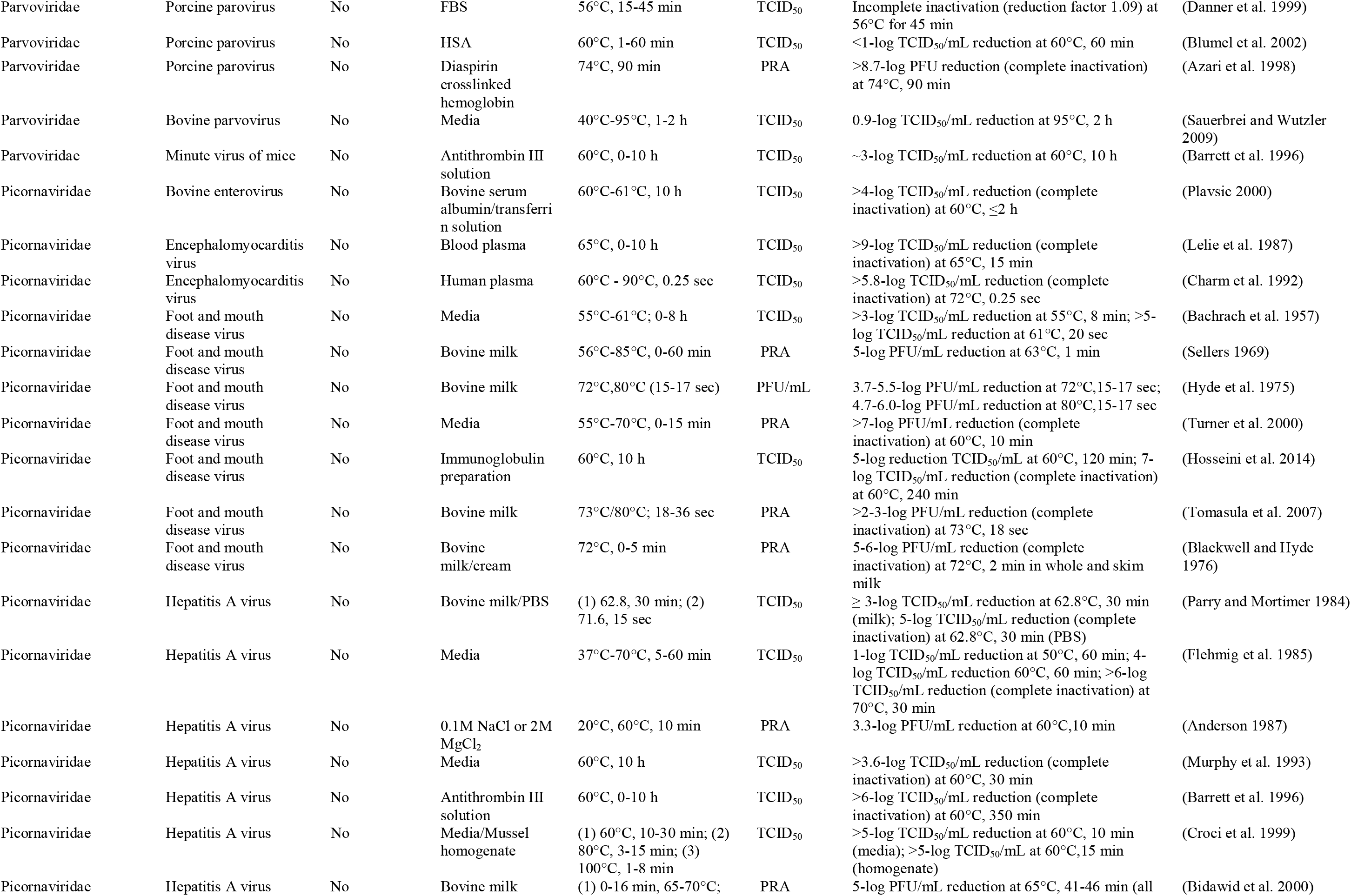
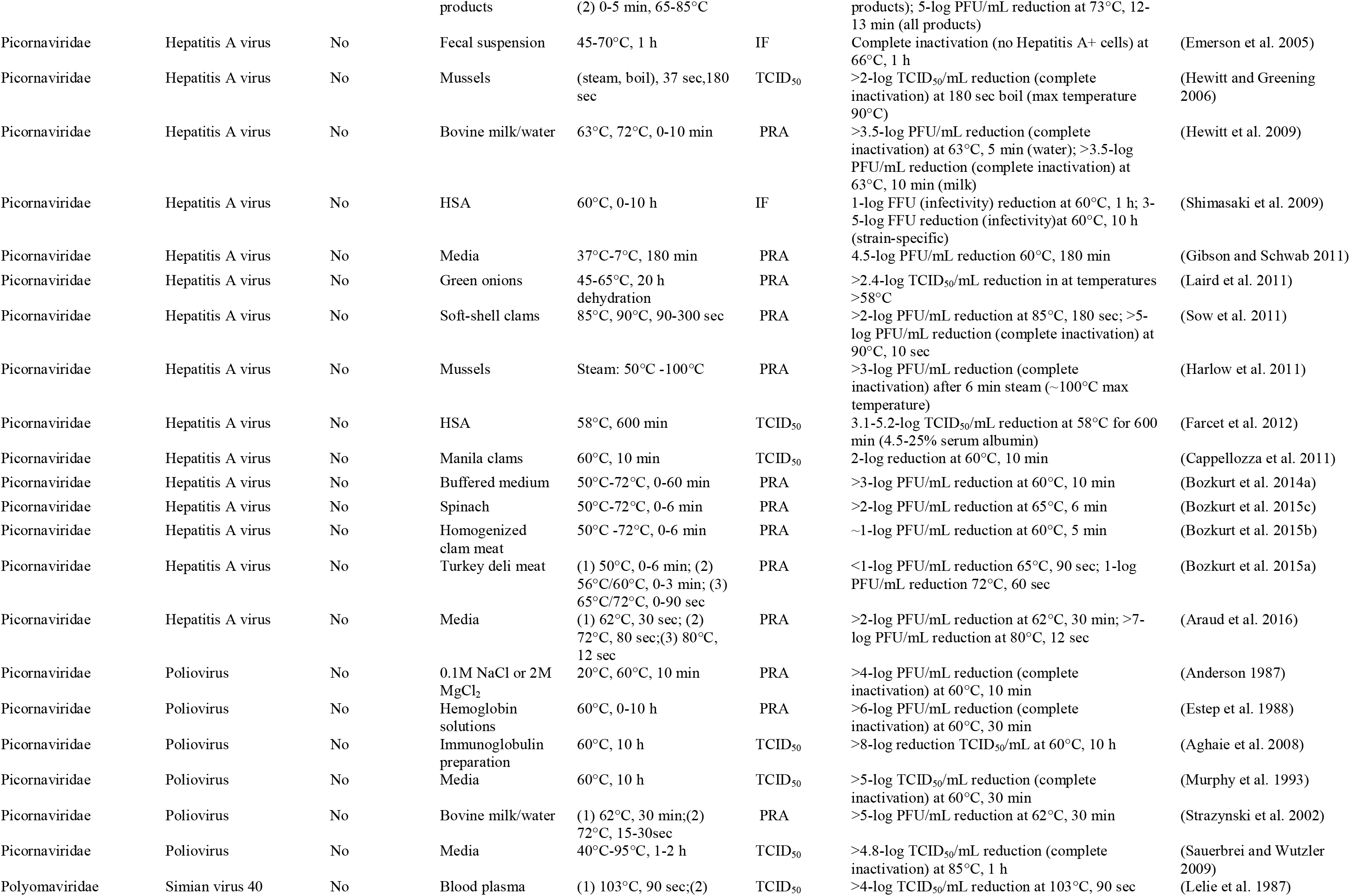
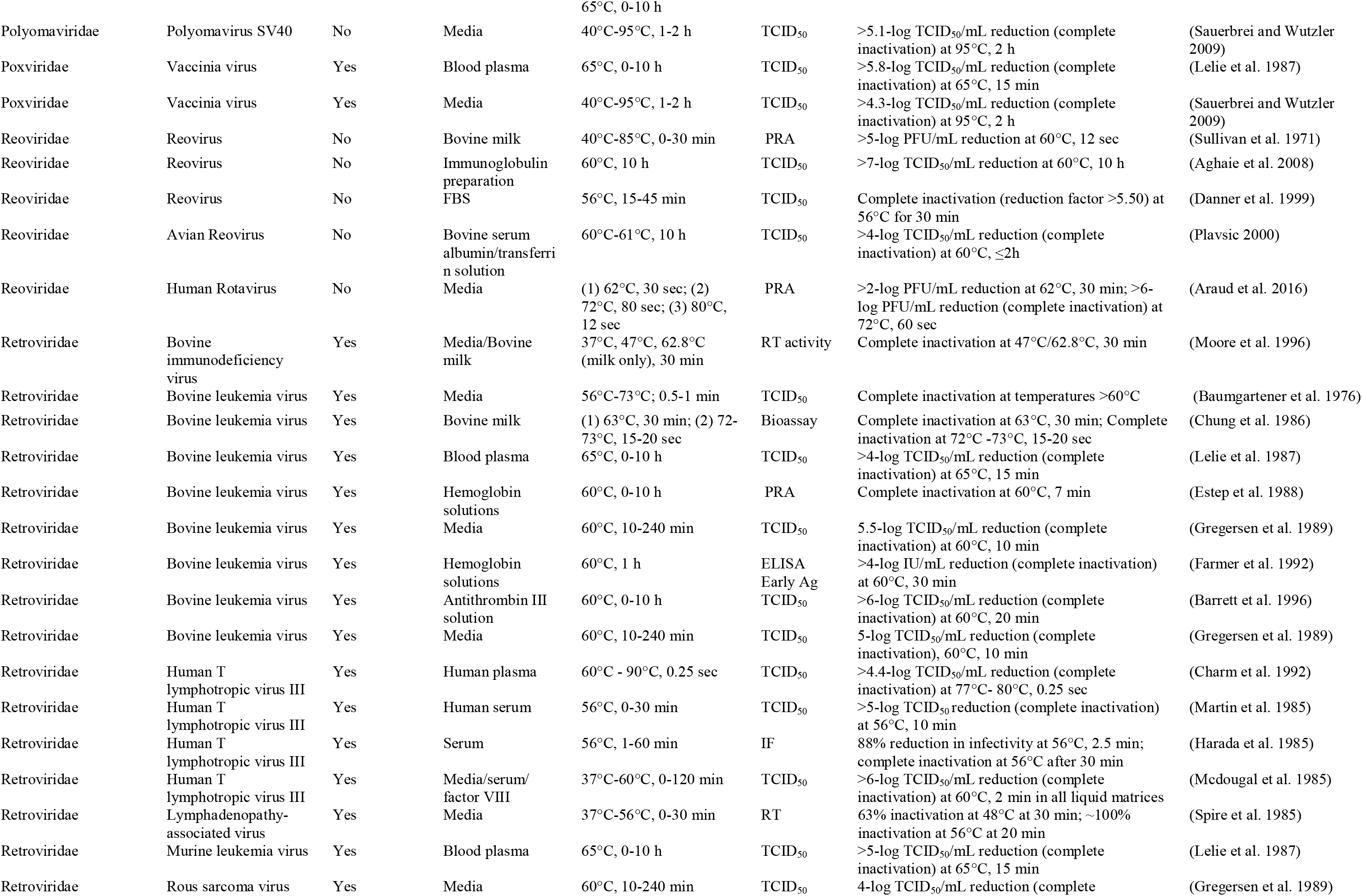
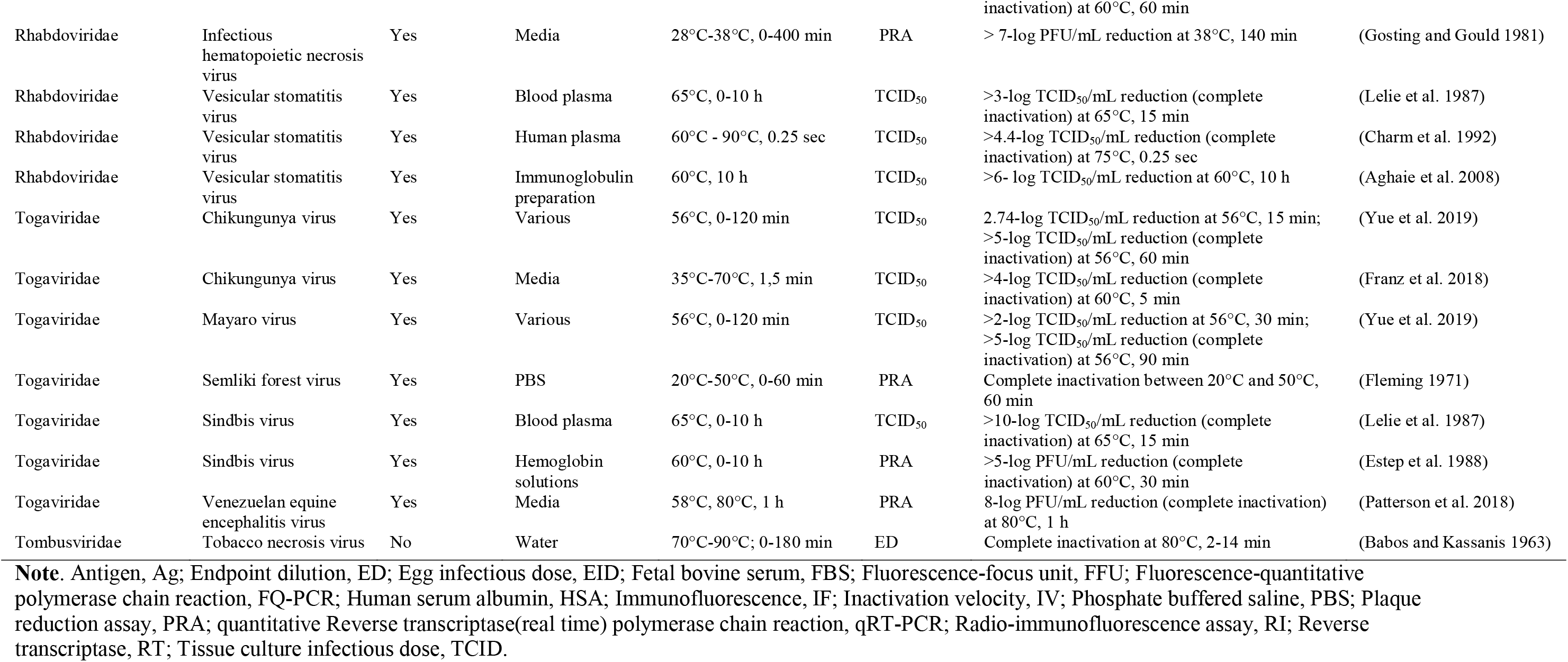
Summary of studies assessing the effect of heat on viral inactivation in matrices others than human milk

Hepatitis A was the most commonly tested virus tested of the picornaviridae family and was seen to be particularly heat sensitive in a variety of matrices including bovine milk, cell culture media and soft-shell clams. For example, a minimum of a 4-log reduction in infectivity of Hepatitis A was observed after different thermal pasteurization parameters such as 60°C-65°C for 10-180 min(Croci et al. 1999; Bidawid et al. 2000; Gibson and Schwab 2011); to 72°C for 1-13 min (Bidawid et al. 2000; Araud et al. 2016), and to 90°C for 5 min (Sow et al. 2011). Murine norovirus, the most frequently tested virus of the caliciviridae family, was also observed to be sensitive to heat. A reduction in infectivity of greater than 5-log was observed at temperatures of 60°C −67°C for 1-60 min (Gibson and Schwab 2011; Shao et al. 2018), >3.5-log reduction at 72°C for 1 min (Hewitt et al. 2009; Araud et al. 2016), and >5-log reduction at 85°C −90°C for 1-5 min (Sow et al. 2011; Park et al. 2014a). HIV-1 was the most commonly tested of the retroviridae and was also susceptible to heat-inactivation. Greater than 4-log reduction in TCID_50_ was observed at 60°C-65°C for 10-15 min (Lelie et al. 1987; Gregersen et al. 1989); similar reductions were observed at 77°C-80°C after 0.25 seconds(Charm et al. 1992). Notably, viruses in the coronaviridae family, SARS-CoV, and SARS-CoV-2, also show significant reductions in infectivity (>5-7-log reduction in TCID_50_/mL) following pasteurization; complete inactivation was observed at temperatures between 56°C-60°C for a 5-60 min duration(Duan et al. 2003; Darnell et al. 2004; Yunoki et al. 2004; Kariwa et al. 2006). Other coronaviruses, including canine coronavirus, and MERS show sensitivities to heating in cell culture media, where a clinically relevant reduction in infectivity (>4.5 – 5.5 log TCID_50_) is attainable upon heating at 65°C for 5-15 minutes(Pratelli 2008; Leclercq et al. 2014). Furthermore, cytomegalovirus, a member of the herpesviridae family was completely inactivated at temperatures between 50°C-65°C for 15-30 min(Plummer and Lewis 1965; Lelie et al. 1987; Farmer et al. 1992; Mikawa et al. 2019).

### Viruses tested in human milk and other matrices

Finally, the summary of the comparisons among viruses that were tested in both a human milk and a non-human milk matrix is shown in Table 3. Overall, the range of temperatures that yielded some degree of log reduction were consistent among viruses, irrespective of the matrix. Cytomegalovirus, for example, was a virus where there was good agreement among studies testing thermal pasteurization in either a human milk or a non-human milk matrix; inactivation was evident at temperatures between 50°C and 65°C for 10-30 min. Similarly consistent, porcine parvovirus in the parvoviridae family was found to be heat resistant in either human milk or non-human milk matrices(Danner et al. 1999; Terpstra et al. 2007; Sauerbrei and Wutzler 2009). There were some differences in the time required for the log reduction in infectivity depending on matrix, but there were no discernable trends identified.

**Table 3.** Comparing the log reductions in detectable live viruses pasteurized in both a human milk and a non-human milk matrix

| Human Milk Matrix | Non-Human Milk Matrix |
| --- | --- |

| Family | Virus | Reduction * | Temperature † | Time ‡ | Reference | Reduction * | Temperature † | Time ‡ | Reference |
| --- | --- | --- | --- | --- | --- | --- | --- | --- | --- |
| Caliciviridae | Feline Calicivirus | >5 | 65 | 2 | (Topping et al. 2009) | 4.5 - >5 | 60 | 1-60 | (Gibson and Schwab 2011; Bozkurt et al. 2014b, 2014a) |
| Flaviviridae | Bovine viral diarrhea virus | ~4 | 72 | 0.1 | (Terpstra et al. 2007) | >6.5 | 60 | <120 | (Plavsic 2000; Aghaie et al. 2008) |
| Flaviviridae | Zika virus | >6 | 63 | 30 | (Hamilton Spence et al. 2017) | >4 | 56-58 | 5-10 | (Blümel et al. 2017; Farcet and Kreil 2017) |
| Herpesviridae | Cytomegalovirus | >3/UD | 56-63 | 8-30 | (Welsh et al. 1979; Friis and Andersen 1982; Goldblum et al. 1984) | >5/UD | 50-65 | 15-30 | (Plummer and Lewis 1965; Lelie et al. 1987; Farmer et al. 1992; Mikawa et al. 2019) |
| Herpesviridae | Herpes Simplex Virus | 4.2 | 63 | 30 | (Welsh et al. 1979) | >3 - >5 | 50-60 | <1-600 | (Plummer and Lewis 1965; Sullivan et al. 1971; Aghaie et al. 2008) |
| Picornaviridae | Hepatitis A Virus | 2 | 72 | 0.3 | (Terpstra et al. 2007) | >3 - >5 | 60-63 | 10-60 | (Parry and Mortimer 1984; Flehmig et al. 1985; Anderson 1987; Murphy et al. 1993; Croci et al. 1999; Bidawid et al. 2000; Araud et al. 2016) |
| Parvoviridae | Porcine Parvovirus | 0.5 | 72 | 0.3 | (Terpstra et al. 2007) | <1 | 56-60 | 15-60 | (Danner et al. 1999; Blumel et al. 2002) |
| Retroviridae | HIV-1 | >5.5 | 62.5 | 30 | (Orloff et al. 1993) | >4 | 60-65 | 7-30 | (Lelie et al. 1987; Estep et al. 1988; Gregersen et al. 1989; Farmer et al. 1992) |
| Togaviridae | Semliki forest virus | 3.2 | 63 | 30 | (Welsh et al. 1979) | UD | 20-50 | 60 | (Fleming 1971) |

**Note.** Undetectable, UD
\*Log-PFU or TCID<sub>50</sub>/mL
† Degree °C
‡ Minutes

## Discussion

Pasteurization is an essential part of human donor milk banking and is practiced worldwide to reduce or eliminate the risk of transmission of viruses that may be expressed in milk or may be found as a contaminant; Holder pasteurization (62.5°C, 30 min) is the most common method used(*Guidelines for the Establishment and Operation of a Donor Human Milk Bank* 2018). Our rapid review aimed to summarize the literature pertaining to the effect of thermal pasteurization on viral load and detectable live virus; in particular, research that has been conducted using a human milk matrix. Our rapid review also aimed to compare viruses that have been both tested in a human milk matrix and a non-human milk matrix to better understand any potential modulating effects.

As expected, the most commonly studied viruses in human milk in relation to thermal pasteurization included those that have been previously shown to be transmitted through breastmilk; primarily cytomegalovirus and HIV-1 which are enveloped viruses belonging to the herpesviridae and retroviridae families respectively(Prendergast et al. 2019). Although not as common as cytomegalovirus or HIV, Ebola, Marburg, and Zika viruses have also been studied in human milk given that viral nucleic acid has been detected in milk and transmission is a potential concern (Hamilton Spence et al. 2017; Sampieri and Montero 2019). Despite differences in viral taxonomy and caspid envelope, pasteurization is effective at significantly reducing detectable virus or viral load by several log, and in many cases, to below detectable levels (Table 1).

Many studies involving human milk tested pasteurization parameters that included the Holder method (62.5°C, 30 minutes) in order to mimic practices at milk banks; however, a variety of time and temperature combinations were tested. Although many studies reported that viruses including Ebola, Marburg, Zika, cytomegalovirus, and HIV appear to be completely inactivated after 30 min at 62.5°C −63°C (Table 1), others report inactivation after a shorter duration; it remains unclear whether Holder pasteurization for shorter times might effectively inactivate these viruses. Arriving at a consensus is difficult given that one study might assess reductions in surviving virus concentrations before and after Holder pasteurization and another might assess at different time points during the pasteurization process. Moreover, high-temperature short-time pasteurization, defined here as pasteurization above 70°C for less than 30 minutes, appears to be as effective as pasteurization at lower temperatures for a longer duration.

Given the limited research in a human milk matrix, the inclusion of studies that assessed viral load or detectable live virus in a range of matrices allowed us to assess a broader scope of viruses belonging to numerous taxonomic families. The matrix may influence the effectiveness of pasteurization by altering how heat is distributed; however, our results suggest that irrespective of matrix, enveloped, compared to non-enveloped viruses, generally require less input of thermal energy in order to achieve similar reductions in viral load or live virus concentration. This suggests that the results presented in Table 2 may, to a certain degree, be representative of how viruses could be inactivated by heat in human milk. In all matrices, including human milk, pasteurization at temperatures of 62.5°C was generally sufficient to reduce surviving viral load by several logs or to below the limit of detection—depending on the starting concentration of virus and whether it was enveloped. To completely inactivate non-enveloped viruses, such as bovine viral diarrhea virus, hepatitis A or porcine parvovirus in human milk or in other matrices, temperatures above 63°C (70°C −90°C) or a significantly longer duration at 60°C-63°C (Table 2) is generally required. Overall, the results are consistent with the logarithmic thermal death time curve where the same degree of thermal lethality can occur at varying temperatures depending on holding time; pasteurization at higher temperatures for shorter durations or lower temperatures for longer durations yielded similar results in terms of the magnitude of infectivity reductions.

Finally, while we cannot discount any differences in response to thermal pasteurization, viruses that were tested in both a human milk and non-human milk matrix appeared to require similar temperatures to elicit a given log reduction in infectivity. Nevertheless, there was significant variability in the duration of pasteurization tested, making it difficult to draw any conclusions as some viruses may require greater time at temperature for one matrix, and less time at temperature for another. In addition to there being a wide range of matrices included as part of the non-human milk group, differences in the time may be an artefact of the design of the respective studies; in many cases, viral infectivity or load was not always assessed longitudinally, but after a predetermined length of time. Consequently, this may overestimate the amount of time required to achieve a certain degree of inactivation, making it difficult to compare and aggregate the results from different studies.

There are many strengths of this rapid review. First, we carried out a robust search strategy, in addition to manual searches of grey literature, to generate a complete list of studies, irrespective of language or year published that assessed the impact of thermal pasteurization on viral load in human milk and other matrices. The studies in this review reported on a wide range of thermal pasteurization parameters (low-temperature long-time, high-temperature short-time) across several viruses in a diverse set of matrices. Despite these, the interpretation of our results should be considered alongside its limitations. First, this review was conducted by a single reviewer which may have introduced potential selection bias during initial screening. As a result, our review may not have captured all possible studies. Despite this, the purpose of this review was to rapidly and broadly characterize how viruses in any matrix, including human milk, might respond to thermal pasteurization. Second, the reduction in viral load or detectable live virus that was extracted was approximated if multiple strains of a given virus genus were studied, despite the potential of strain-specific variation in thermal resistance. Third, in our comparison of studies that assessed similar viruses in both a human milk and non-human milk matrix, we chose to aggregate the results to match, to the best of our ability, the pasteurization parameters tested in human milk. While this may have allowed us to assess the temperature and time requirements to achieve a certain log reduction, we were limited to a narrow range of pasteurization conditions.

To our knowledge, this rapid review is the first to broadly summarize the literature that has reported on the impact of any thermal pasteurization on virus survival. The results from this study highlight our limited understanding with respect to the effect of thermal pasteurization on viruses in human milk— this is especially relevant given the possibility that novel viruses, including SARS-CoV-2, may be present in human milk. Although currently, there is insufficient evidence to suggest that SARS-CoV-2 is expressed in milk and could lead to vertical transmission, it may also be present as a contaminant (Lackey et al. 2020). Based on the literature review, Holder pasteurization (62.5°C, 30 minutes) may be sufficient to inactivate non-heat resistant viruses that may be present in HM, including coronaviruses. Though our attempt to rapidly survey all known viral families may help provide some insight into how novel viruses may respond to thermal pasteurization, additional research is warranted to synthesize empirical evidence using human milk as the matrix.

## Data Availability

Data sharing is not applicable to this article as no datasets were generated or analyzed during the current study.

## Acknowledgments

This work was supported by the Ontario Graduate Scholarship; Restracomp Scholarship, The Hospital for Sick Children and the Canadian Institutes of Health Research [FDN# 143233] The authors gratefully acknowledge Glyneva Bradley-Ridout at the University of Toronto who was consulted on the search strategy. All authors have no conflicts of interest to disclose.

## References

Adcock, W.L., MacGregor, A., Davies, J.R., Hattarki, M., Anderson, D.A., and Goss, N.H. 1998. Chromatographic removal and heat inactivation of hepatitis B virus during the manufacture of human albumin. Biotechnol. Appl. Biochem. 28 (Pt2): 169–78. doi:10.1111/j.1470-8744.1998.tb00516.x.

Aghaie, A., Pourfatollah, A.A., Bathaie, S.Z., Moazzeni, S.M., Pour, H.K.M., and Sharifi, Z. 2008. Inactivation of virus in intravenous immunoglobulin G using solvent/detergent treatment and pasteurization. Hum. Antibodies 17(3–4): 79–84. doi:10.3233/hab-2008-173-405.

Ailavadi, S., Davidson, P.M., Morgan, M.T., and D’Souza, D.H. 2019. Thermal Inactivation Kinetics of Tulane Virus in Cell-Culture Medium and Spinach. J. Food Sci. 84(3): 557–563. doi:10.1111/1750-3841.14461.

Alexander, D.J., and Manvell, R.J. 2004. Heat inactivation of Newcastle disease virus (strain Herts 33/56) in artificially infected chicken meat homogenate. Avian Pathol. 33(2): 222–225. doi:10.1080/0307945042000195795.

Anderson, D.A. 1987. Cytopathology, plaque assay, and heat inactivation of hepatitis A virus strain HM175. J. Med. Virol. 22(1): 35-44. doi:10.1002/jmv.1890220106.

Araud, E., DiCaprio, E., Ma, Y., Lou, F., Gao, Y., Kingsley, D., Hughes, J.H., and Li, J. 2016. Thermal inactivation of enteric viruses and bioaccumulation of enteric foodborne viruses in live oysters (Crassostrea virginica). Appl. Environ. Microbiol. 82(7): 2086–2099. doi:10.1128/AEM.03573-15.

Arita, M., and Matumoto, M. 1968. Heat Inactivation of Measles Virus. Jpn. J. Microbiol. 12(1): 121–122. doi:10.1111/j.1348-0421.1968.tb00374.x.

Arslanoglu, S., Bertino, E., Tonetto, P., De Nisi, G., Ambruzzi, A.M., Biasini, A., Profeti, C., Spreghini, M.R., and Moro, G.E. 2010. Guidelines for the establishment and operation of a donor human milk bank. J. Matern. Neonatal Med. 23(sup2): 1–20. doi:10.3109/14767058.2010.512414.

Azari, M., Ebeling, A., Baker, R., Burhop, K., Camacho, T., Estep, T., Guzder, S., Marshall, T., Rohn, K., Sarajari, R., Boose, J.A., Gauvin, G., Homer, R., Lu, B., Pearson, L., and Vacante, D. 1998. Validation of the Heat Treatment Step Used in the Production of Diaspirln Crosslinked Hemoglobin (DCLHb^TM^) For Viral Inactivation. Artif. Cells, Blood Substitutes, Biotechnol. 26(5–6): 577–582. doi:10.3109/10731199809117477.

Babos, P., and Kassanis, B. 1963. Thermal inactivation of tobacco necrosis virus. Virology 20(3): 490–497. doi:10.1016/0042-6822(63)90099-0.

Bachrach, H.L., Breese, S.S., Callis, J.J., Hess, W.R., and Patty, R.E. 1957. Inactivation of Foot-and-Mouth Disease Virus by pH and Temperature Changes and by Formaldehyde. Exp. Biol. Med. 95(1): 147–152. doi:10.3181/00379727-95-23148.

Baert, L., Uyttendaele, M., Van Coillie, E., and Debevere, J. 2008. The reduction of murine norovirus 1, B. fragilis HSP40 infecting phage B40–8 and E. coli after a mild thermal pasteurization process of raspberry puree. Food Microbiol. 25(7): 871–874.doi:10.1016/j.fm.2008.06.002.

Barnaud, E., Rogée, S., Garry, P., Rose, N., and Pavio, N. 2012. Thermal inactivation of infectious hepatitis E virus in experimentally contaminated food. Appl. Environ. Microbiol. 78(15): 5153–5159. doi:10.1128/AEM.00436-12.

Barrett, P.N., Meyer, H., Wachtel, I., Eibl, J., and Dorner, F. 1996. Determination of the inactivation kinetics of hepatitis A virus in human plasma products using a simple TCID50 assay. J. Med. Virol. 49(1): 1–6. doi:10.1002/(SICI)1096-071(199605)49:1<1::AID-JMV1>3.0.CO;2-A.

Baumgartener, L., Olson, C., and Onuma, M. 1976. Effect of Pasteurization and Heat Treatment on Bovine Leukemia Virus. J Am Vet Med Assoc 169(11): 1189–91.

Bidawid, S., Farber, J.M., Sattar, S.A., and Hayward, S. 2000. Heat inactivation of hepatitis A virus in dairy foods. J. Food Prot. 63(4): 522–528. doi:10.4315/0362-028X-63.4.522.

Blackwell, J.H., and Hyde, J.L. 1976. Effect of heat on foot-and-mouth disease virus (FMDV) in the components of milk from FMDV-infected cows. J. Hyg. (Lond). 77(1): 77–83. doi:10.1017/s0022172400055534.

Blümel, J., Musso, D., Teitz, S., Miyabayashi, T., Boller, K., Schnierle, B.S., and Baylis, S.A. 2017. Inactivation and removal of Zika virus during manufacture of plasma-derived medicinal products. Transfusion 57(3): 790–796. doi:10.1111/trf.13873.

Blumel, J., Schmidt, I., Willkommen, H., and Lower, J. 2002. Inactivation of parvovirus B19 during pasteurization of human serum albumin. Transfusion 42(8): 1011–1018. doi:10.1046/j.1537-2995.2002.00158.x.

Bozkurt, H., D’Souza, D.H., and Davidson, P.M. 2014a. A comparison of the thermal inactivation kinetics of human norovirus surrogates and hepatitis A virus in buffered cell culture medium. Food Microbiol. 42: 212–217. Elsevier Ltd. doi:10.1016/j.fm.2014.04.002.

Bozkurt, H., D’Souza, D.H., and Davidson, P.M. 2015a. Thermal inactivation kinetics of human norovirus surrogates and hepatitis A virus in turkey deli meat. Appl. Environ. Microbiol. 81(14): 4850–4859. doi:10.1128/AEM.00874-15.

Bozkurt, H., D’Souza, D.H., and Davidson, P.M. 2015b. Thermal inactivation kinetics of hepatitis A virus in homogenized clam meat (Mercenaria mercenaria). J. Appl. Microbiol. 119(3): 834–844. doi:10.1111/jam.12892.

Bozkurt, H., Leiser, S., Davidson, P.M., and D’Souza, D.H. 2014b. Thermal inactivation kinetic modeling of human norovirus surrogates in blue mussel (Mytilus edulis) homogenate. Int. J. Food Microbiol. 172: 130–136. Elsevier B.V. doi:10.1016/j.ijfoodmicro.2013.11.026.

Bozkurt, H., Ye, X., Harte, F., D’Souza, D.H., and Davidson, P.M. 2015c. Thermal inactivation kinetics of hepatitis A virus in spinach. Int. J. Food Microbiol. 193: 147–151. Elsevier B.V. doi:10.1016/j.ijfoodmicro.2014.10.015.

Cappellozza, E., Arcangeli, G., Rosteghin, M., Sulaj, K., Magnabosco, C., Bertoli, E., and Terregino, C. 2011. Survival of hepatitis a virus in pasteurized manila clams. Ital. J. Food Sci. 24(3): 247–253.

Charm, S.E., Landau, S., Williams, B., Horowitz, B., Prince, A.M., and Pascual, D. 1992. High-Temperature Short-Time Heat Inactivation of HIV and Other Viruses in Human Blood Plasma. Vox Sang. 62(1): 12–20. doi:10.1111/j.1423-0410.1992.tb01160.x.

Chin, A.W.H., Chu, J.T.S., Perera, M.R.A., Hui, K.P.Y., Yen, H.-L., Chan, M.C.W., Peiris, M., and Poon, L.L.M. 2020. Stability of SARS-CoV-2 in different environmental conditions. The Lancet Microbe: 0–4. doi:10.1016/s2666-5247(20)30003-3.

Chmielewski, R.A., Beck, J.R., and Swayne, D.E. 2011. Thermal inactivation of avian influenza virus and newcastle disease virus in a fat-free egg product. J. Food Prot. 74(7): 1161–1168. doi:10.4315/0362-028X.JFP-10-415.

Chu, C.M. 1948. Inactivation of haemagglutinin and infectivity of influenza and Newcastle disease viruses by heat and by formalin. J. Hyg. (Lond). 46(3): 247–251. doi:10.1017/S0022172400036366.

Chung, Y.S., Prior, H.C., Duffy, P.F., Rogers, R.J., and Mackenzie, A.R. 1986. The effect of pasteurisation on bovine leucosis virus-infected milk. Aust. Vet. J. 63(11): 379-380. doi:10.1111/j.1751-0813.1986.tb02908.x.

COVID-19: EMBA Position Statement. 2020.

Croci, L., Ciccozzi, M., De Medici, D., Di Pasquale, S., Fiore, A., Mele, A., and Toti, L. 1999. Inactivation of Hepatitis A virus in heat-treated mussels. J. Appl. Microbiol. 87(6): 884–888. doi:10.1046/j.1365-2672.1999.00935.x.

Danner, D.J., Smith, J., and Plavsic, M. 1999. Inactivation of viruses and mycoplasmas in fetal bovine serum using 56°C heat. BioPharm 12(6): 50–52.

Darnell, M.E.R., Subbarao, K., Feinstone, S.M., and Taylor, D.R. 2004. Inactivation of the coronavirus that induces severe acute respiratory syndrome, SARS-CoV. J. Virol. Methods 121(1): 85–91. doi:10.1016/j.jviromet.2004.06.006.

Darnell, M.E.R., and Taylor, D.R. 2006. Evaluation of inactivation methods for severe acute respiratory syndrome coronavirus in noncellular blood products. Transfusion 46(10): 1770–1777. doi:10.1111/j.1537-2995.2006.00976.x.

Donalisio, M., Cagno, V., Vallino, M., Moro, G.E., Arslanoglu, S., Tonetto, P., Bertino, E., and Lembo, D. 2014. Inactivation of high-risk human papillomaviruses by Holder pasteurization: implications for donor human milk banking. J. Perinat. Med. 42(1): 1–8. doi:10.1515/jpm-2013-0200.

Duan, S.M., Zhao, X.S., Wen, R.F., Huang, J.J., Pi, G.H., Zhang, S.X., Han, J., Bi, S.L., Ruan, L., and Dong, X.P. 2003. Stability of SARS Coronavirus in Human Specimens and Environment and Its Sensitivity to Heating and UV Irradiation. Biomed. Environ. Sci. 16(3): 246–255.

Duizer, E., Bijkerk, P., Rockx, B., de Groot, A., Twisk, F., and Koopmans, M. 2004. Inactivation of Caliciviruses. Appl. Environ. Microbiol. 70(8): 4538–4543. doi:10.1128/AEM.70.8.4538-4543.2004.

Dworsky, M., Stagno, S., Pass, R.F., Cassady, G., and Alford, C. 1982. Persistence of cytomegalovirus in human milk after storage. J. Pediatr. 101(3): 440–443. doi:10.1016/S0022-3476(82)80081-4.

Eglin, RP, Wilkinson, A. 1987. HIV Infection and Pasteurization of Breast Milk--Letter to Editor. Lancet: 1093.

Emerson, S.U., Arankalle, V.A., and Purcell, R.H. 2005. Thermal Stability of Hepatitis E Virus. J. Infect. Dis. 192(5): 930–933. doi:10.1086/432488.

Estep, T.N., Bechtel, M.K., Miller, T.J., and Bagdasarian, A. 1988. Virus inactivation in hemoglobin solutions by heat. Biomater. Artif. Cells Artif. Organs 16(3--Jan): 129–134. doi:10.3109/10731198809132563.

Farcet, M.R., Kindermann, J., Modrof, J., and Kreil, T.R. 2012. Inactivation of hepatitis A variants during heat treatment (pasteurization) of human serum albumin. Transfusion 52(1): 181–187. doi:10.1111/j.1537-2995.2011.03251.x.

Farcet, M.R., and Kreil, T.R. 2017. Zika virus is not thermostable: very effective virus inactivation during heat treatment (pasteurization) of human serum albumin. Transfusion 57(3): 797–801. doi:10.1111/trf.13953.

Farmer, M., Ebeling, A., Marshall, T., Hauck, W., Sun, C.-S., White, E., and Long, Z. 1992. Validation of Virus Inactivation by Heat Treatment in the Manufacture of Diaspqun Crosslinksd Hemoglobin. Biomater. Artif. Cells Immobil. Biotechnol. 20(2–4): 429–433. doi:10.3109/10731199209119663.

Flehmig, B., Billing, A., Vallbracht, A., and Botzenhart, K. 1985. Inactivation of hepatitis A virus by heat and formaldehyde. Water Sci. Technol. 17(10): 43–45. doi:10.2166/wst.1985.0094.

Fleming, P. 1971. Thermal inactivation of Semliki Forest virus. J. Gen. Virol. 13(3): 385–391. doi:10.1099/0022-1317-13-3-385.

De Flora, S., and Badolati, G. 1973. Thermal inactivation of untreated and gamma irradiated A2/Aichi/2/68 influenza virus. J. Gen. Virol. 20(2): 261–265. doi:10.1099/0022-1317-20-2-261.

Franz, S., Friesland, M., Passos, V., Todt, D., Simmons, G., Goffinet, C., and Steinmann, E. 2018. Susceptibility of Chikungunya Virus to Inactivation by Heat and Commercially and World Health Organization-Recommended Biocides. J Infect Dis 218(9): 1507–1510. doi:10.1093/infdis/jiy359.

Friis, H., and Andersen, H.K. 1982. Rate of inactivation of cytomegalovirus in raw banked milk during storage at −20°C and pasteurisation. Br. Med. J. 285(6355): 1604–1605. doi:10.1136/bmj.285.6355.1604.

Furlow, B. 2020. US NICUs and donor milk banks brace for COVID-19. Lancet Child Adolesc. Heal. 4(5): 355. doi:10.1016/S2352-4642(20)30103-6.

Gibson, K.E., and Schwab, K.J. 2011. Thermal Inactivation of Human Norovirus Surrogates. Food Environ. Virol. 3(2): 74–77. doi:10.1007/s12560-011-9059-4.

Goldblum, R.M., Dill, C.W., Albrecht, T.B., Alford, E.S., Garza, C., and Goldman, A.S. 1984. Rapid high-temperature treatment of human milk. J. Pediatr. 104(3): 380–385. doi:10.1016/S0022-3476(84)81099-9.

Gosting, L.H., and Gould, R.W. 1981. Thermal inactivation of infectious hematopoietic necrosis and infectious pancreatic necrosis viruses. Appl. Environ. Microbiol. 41(4): 1081–1082. doi:10.1128/aem.41.4.1081-1082.1981.

Gregersen, J.P., Hilfenhaus, J., and Lemp, J.F. 1989. Heat inactivation of human immunodeficiency virus type 2 (HIV-2). J. Biol. Stand. 17(4): 377–379. doi:10.1016/S0092-1157(89)80009-5.

Guidelines for the Establishment and Operation of a Donor Human Milk Bank. 2018. In 10th Editi. Human Milk Association of North America, Fort Worth, TX.

Hamilton Spence, E., Huff, M., Shattuck, K., Vickers, A., Yun, N., and Paessler, S. 2017. Ebola Virus and Marburg Virus in Human Milk Are Inactivated by Holder Pasteurization. J. Hum. Lact. 33(2): 351–354. doi:10.1177/0890334416685564.

Hamprecht, K., Maschmann, J., Müller, D., Dietz, K., Besenthal, I., Goelz, R., Middeldorp, J.M., Speer, C.P., and Jahn, G. 2004. Cytomegalovirus (CMV) inactivation in breast milk: Reassessment of pasteurization and freeze-thawing. Pediatr. Res. 56(4): 529–535. doi:10.1203/01.PDR.0000139483.35087.BE.

Harada, S., Yoshiyama, H., and Yamamoto, N. 1985. Effect of heat and fresh human serum on the infectivity of human T-cell lymphotropic virus type III evaluated with new bioassay systems. J. Clin. Microbiol. 22(6): 908–911. doi:10.1128/jcm.22.6.908-911.1985.

Harlow, J., Oudit, D., Hughes, A., and Mattison, K. 2011. Heat Inactivation of Hepatitis A Virus in Shellfish Using Steam. Food Environ. Virol. 3(1): 31–34. doi:10.1007/s12560-010-9052-3.

Hewitt, J., and Greening, G.E. 2006. Effect of heat treatment on hepatitis A virus and norovirus in New Zealand Greenshell mussels (Perna canaliculus) by quantitative real-time reverse transcription PCR and cell culture. J. Food Prot. 69(9): 2217–2223. doi:10.4315/0362-028X-69.9.2217.

Hewitt, J., Rivera-Aban, M., and Greening, G.E. 2009. Evaluation of murine norovirus as a surrogate for human norovirus and hepatitis A virus in heat inactivation studies. J. Appl. Microbiol. 107(1): 65–71. doi:10.1111/j.1365-2672.2009.04179.x.

Hoque, S.A., Hoshino, H., Anwar, K.S., Tanaka, A., Shinagawa, M., Hayakawa, Y., Okitsu, S., Wada, Y., and Ushijima, H. 2013. Transient heating of expressed breast milk up to 65°C inactivates HIV-1 in milk: A simple, rapid, and cost-effective method to prevent postnatal transmission. J. Med. Virol. 85(2): 187–193. doi:10.1002/jmv.23457.

Hosseini, K.M., Dinarvand, R., Rezvan, H., and Jalili, M.A.L.I. 2014. Pasteurization of IgM-enriched Immunoglobulin. (June 2004).

Hyde, J.L., Blackwell, J.H., and Callis, J.J. 1975. Effect of pasteurization and evaporation on foot-and-mouth disease virus in whole milk from infected cows. Can. J. Comp. Med. 39(3): 305–309.

Imagawa, T., Sugiyama, R., Shiota, T., Li, T.C., Yoshizaki, S., Wakita, T., and Ishii, K. 2018. Evaluation of heating conditions for inactivation of hepatitis e virus genotypes 3 and 4. J. Food Prot. 81(6): 947–952. doi:10.4315/0362-028X.JFP-17-290.

Israel-Ballard, K., Donovan, R., Chantry, C., Coutsoudis, A., Sheppard, H., Sibeko, L., and Abrams, B. 2007. Flash-heat inactivation of HIV-1 in human milk: A potential method to reduce postnatal transmission in developing countries. J. Acquir. Immune Defic. Syndr. 45(3): 318–323. doi:10.1097/QAI.0b013e318074eeca.

Jeffery, B.S., Webber, L., Mokhondo, K.R., and Erasmus, D. 2001. Determination of the effectiveness of inactivation of human immunodeficiency virus by Pretoria Pasteurization. J. Trop. Pediatr. 47(6): 345–349. doi:10.1093/tropej/47.6.345.

Kariwa, H., Fujii, N., and Takashima, I. 2006. Inactivation of SARS coronavirus by means of povidone-iodine, physical conditions and chemical reagents. Dermatology 212(SUPPL. 1): 119–123. doi:10.1159/000089211.

King, A.M., Adams, M.J., Carstens, E.B., and Lefkowitz, E.J. (*Editors*). 2012. Virus Taxonomy-Classification and Nomenclature of Viruses-Ninth Report of the International Committee on Taxonoomy of Viruses.

Klotz, D., Schreiner, M., Falcone, V., Jonas, D., Kunze, M., Weber, A., Fuchs, H., and Hentschel, R. 2018. High-temperature short-time treatment of human milk for bacterial count reduction. Front. Pediatr. 6(November): 1–8. doi:10.3389/fped.2018.00359.

Lackey, K., Pace, R., Williams, J., Bode, L., Donovan, S., Järvinen, K., Seppo, A., Raiten, D., Meehan, C., McGuire, M., and McGuire, M. 2020. SARS-CoV-2 and human milk: what is the evidence? Infect. Dis. (Auckl). doi:10.1101/2020.04.07.20056812.

Laird, D.T., Sun, Y., Reineke, K.F., and Carol Shieh, Y. 2011. Effective hepatitis A virus inactivation during low-heat dehydration of contaminated green onions. Food Microbiol. 28(5): 998–1002. Elsevier Ltd. doi:10.1016/j.fm.2011.01.011.

Laude, H. 1981. Thermal inactivation studies of a coronavirus, transmissible gastroenteritis virus. J. Gen. Virol. 56(2): 235–240. doi:10.1099/0022-1317-56-2-235.

Leclercq, I., Batéjat, C., Burguière, A.M., and Manuguerra, J.C. 2014. Heat inactivation of the Middle East respiratory syndrome coronavirus. Influenza Other Respi. Viruses 8(5): 585–586. doi:10.1111/irv.12261.

Lelie, P.N., Reesink, H.W., and Lucas, C.J. 1987. Inactivation of 12 viruses by heating steps applied during manufacture of a hepatitis B vaccine. J. Med. Virol. 23(3): 297–301. doi:10.1002/jmv.1890230313.

Li, X., Huang, R., and Chen, H. 2017. Evaluation of Assays to Quantify Infectious Human Norovirus for Heat and High-Pressure Inactivation Studies Using Tulane Virus. Food Environ. Virol. 9(3): 314–325. Springer US. doi:10.1007/s12560-017-9288-2.

Madani, T.A., Abuelzein, E.T.M.E., Azhar, E.I., and Al-Bar, H.M.S. 2014. Thermal inactivation of Alkhumra hemorrhagic fever virus. Arch. Virol. 159(10): 2687–2691. doi:10.1007/s00705-014-2134-z.

Makhija, A., and Kumar, S. 2017. Characterization of duck plague virus stability at extreme conditions of temperature, pH and salt concentration. Biologicals 45: 102–105. Elsevier Ltd. doi:10.1016/j.biologicals.2016.09.009.

Martin, L.S., McDougal, S.J., and Loskoski, S.L. 1985. Disinfection and Inactivation of the Human T Lymphotropic Virus Type III / Lymphadenopathy-Associated Virus Linda S. Martin, J. Steven McDougal and Sherry L. Loskoski Published by: Oxford University Press Stable URL: https://www.jstor.org/stable/301. J. Infect. Dis. 152(2): 400–403.

Maschmann, J., Müller, D., Lazar, K., Goelz, R., and Hamprecht, K. 2019. New short-term heat inactivation method of cytomegalovirus (CMV) in breast milk: Impact on CMV inactivation, CMV antibodies and enzyme activities. Arch. Dis. Child. Fetal Neonatal Ed. 104(6): F604–F608. doi:10.1136/archdischild-2018-316117.

Mcdougal, J.S., Martin, L.S., Cort, S.P., Mozen, M., Heldebrant, C.M., and Evatt, B.L. 1985. Thermal Inactivation of the Acquired Immunodeficiency Syndrome Virus,. J. Clin. Invest. 76(August): 875–877.

Michie, C.A. 2001. Breast feeding and the risks of viral transmission. Arch. Dis. Child. 84(5): 381–382. doi:10.1136/adc.84.5.381.

Mikawa, T., Mizuno, K., Tanaka, K., Kohda, C., Ishii, Y., Yamamoto, K., and Kobayashi, S. 2019. Microwave treatment of breast milk for prevention of cytomegalovirus infection. Pediatr. Int. 61(12): 1227–1231. doi:10.1111/ped.13954.

Milk Banking and COVID-19. 2020.

Mohandas, S., and Pannaraj, P.S. 2020. Beyond the Bacterial Microbiome: Virome of Human Milk and Effects on the Developing Infant. pp. 1–8. doi:10.1159/000504997.

Moher, D., Liberati, A., Tetzlaff, J., and Altman, D.G. 2009. Preferred Reporting Items for Systematic Reviews and Meta-Analyses: The PRISMA Statement. PLoS Med. 6(7): e1000097. doi:10.1371/journal.pmed.1000097.

Moore, E.C., Keil, D., and St. Cyr Coats, K. 1996. Thermal inactivation of bovine immunodeficiency virus. Appl. Environ. Microbiol. 62(11): 4280–4283. doi:10.1128/aem.62.11.4280-4283.1996.

Murphy, P., Nowak, T., Lemon, S.M., and Hilfenhaus, J. 1993. Inactivation of hepatitis a virus by heat treatment in aqueous solution. J. Med. Virol. 41(1): 61–64. doi:10.1002/jmv.1890410113.

Natal, G. 2019. Searching the Grey Literature: A Handbook for Searching Reports, Working Papers, and Other Unpublished Research. J. Med. Libr. Assoc. 107(2): 276. doi:10.5195/jmla.2019.640.

Orloff, S.L., Wallingford, J.C., and McDougal, J.S. 1993. Inactivation of Human Immunodeficiency Virus Type I in Human Milk: Effects of Intrinsic Factors in Human Milk and of Pasteurization. J. Hum. Lact. 9(1): 13–17. doi:10.1177/089033449300900125.

Park, S.Y., Bae, S.C., and Ha, S. Do. 2014a. Heat Inactivation of a Norovirus Surrogate in Cell Culture Lysate, Abalone Meat, and Abalone Viscera. Food Environ. Virol. 7(1): 58–66. doi:10.1007/s12560-014-9176-y.

Park, S.Y., Kim, S.H., Ju, I.S., Cho, J.I., and Ha, S.D. 2014b. Thermal Inactivation of Murine Norovirus-1 in Suspension and in Dried Mussels (Mytilus edulis). J. Food Saf. 34(3): 193–198. doi:10.1111/jfs.12113.

Parry, J. V., and Mortimer, P.P. 1984. The heat sensitivity of hepatitis A virus determined by a simple tissue culture method. J. Med. Virol. 14(3): 277–283. doi:10.1002/jmv.1890140312.

Patterson, E.I., Warmbrod, K.L., Bouyer, D.H., and Forrester, N.L. 2018. Evaluation of the inactivation of Venezuelan equine encephalitis virus by several common methods. J. Virol. Methods 254(January): 31–34. Elsevier. doi:10.1016/j.jviromet.2018.01.009.

Pfaender, S., Vielle, N.J., Ebert, N., Steinmann, E., Alves, M.P., and Thiel, V. 2017. Inactivation of Zika virus in human breast milk by prolonged storage or pasteurization. Virus Res. 228: 58–60. Elsevier B.V. doi:10.1016/j.virusres.2016.11.025.

Plavsic, Z.M. 2000. Effect of heat treatment on four viruses inoculated into BSA and bovine transferrin solution. BioPharm 13(6): 54–56.

Plummer, G., and Lewis, B. 1965. Thermoinactivation of Herpes Simplex Virus and Cytomegalovirus. J. Bacteriol. 89(3): 671–674. doi:10.1128/jb.89.3.671-674.1965.

Pratelli, A. 2008. Canine coronavirus inactivation with physical and chemical agents. Vet. J. 177(1): 71–79. doi:10.1016/j.tvjl.2007.03.019.

Prendergast, A.J., Goga, A.E., Waitt, C., Gessain, A., Taylor, G.P., Rollins, N., Abrams, E.J., Lyall, E.H., and de Perre, P. Van. 2019. Transmission of CMV, HTLV-1, and HIV through breastmilk. Lancet Child Adolesc. Heal. 3(4): 264-273. doi:10.1016/S2352-4642(19)30024-0.

Quigley, M., Embleton, N.D., and McGuire, W. 2019. Formula versus donor breast milk for feeding preterm or low birth weight infants. Cochrane database Syst. Rev. 7: CD002971. doi:10.1002/14651858.CD002971.pub5.

Sampieri, C.L., and Montero, H. 2019. Breastfeeding in the time of Zika: a systematic literature review. PeerJ 7: e6452. doi:10.7717/peerj.6452.

Sauerbrei, A., and Wutzler, P. 2009. Testing thermal resistance of viruses. Arch. Virol. 154(1): 115–119. doi:10.1007/s00705-008-0264-x.

Sellers, R.F. 1969. Inactivation of foot-and-mouth disease virus in milk. Br. Vet. J. 125(4): 163–168. Elsevier Masson SAS. doi:10.1016/S0007-1935(17)49008-7.

Shao, L., Chen, H., Hicks, D., and Wu, C. 2018. Thermal inactivation of human norovirus surrogates in oyster homogenate. Int. J. Food Microbiol. 281(October 2017): 47–53. Elsevier. doi:10.1016/j.ijfoodmicro.2018.05.013.

Shimasaki, N., Kiyohara, T., Totsuka, A., Nojima, K., Okada, Y., Yamaguchi, K., Kajioka, J., Wakita, T., and Yoneyama, T. 2009. Inactivation of hepatitis A virus by heat and high hydrostatic pressure: Variation among laboratory strains. Vox Sang. 96(1): 14–19. doi:10.1111/j.1423-0410.2008.01113.x.

Song, H., Li, J., Shi, S., Yan, L., Zhuang, H., and Li, K. 2010. Thermal stability and inactivation of hepatitis C virus grown in cell culture. Virol. J. 7: 1–12. doi:10.1186/1743-422X-7-40.

Song, X., Ju, L., Wei, G., and Jian, Q. 2011. Heat Impact Upon the Infectivity of Hepatitis B Virus in Serum. Zhonghua Yu Fang Yi Xue Za Zhi 45(8): 723–6.

Sow, H., Desbiens, M., Morales-Rayas, R., Ngazoa, S.E., and Jean, J. 2011. Heat inactivation of hepatitis A virus and a norovirus surrogate in soft-shell clams (Mya arenaria). Foodborne Pathog. Dis. 8(3): 387–393. doi:10.1089/fpd.2010.0681.

Spire, B., Barré-Sinoussi, F., Dormont, D., Montagnier, L., and Chermann, J.C. 1985. Inactivation of Lymphadenopathy-Associated Virus By Heat, Gamma Rays, and Ultraviolet Light. Lancet 325(8422): 188–189. doi:10.1016/S0140-6736(85)92026-4.

Strazynski, M., Krämer, J., and Becker, B. 2002. Thermal inactivation of poliovirus type 1 in water, milk and yoghurt. Int. J. Food Microbiol. 74(1–2): 73–78. doi:10.1016/S0168-1605(01)00708-5.

Sullivan, R., Tierney, J.T., Larkin, E.P., Read, R.B., and Peeler, J.T. 1971. Thermal resistance of certain oncogenic viruses suspended in milk and milk products. Appl. Microbiol. 22(3): 315–320. doi:10.1128/AEM.22.3.315-320.1971.

Terpstra, F.G., Rechtman, D.J., Lee, M.L., Van Hoeij, K., Berg, H., Van Engelenberg, F.A.C., and Van’t Wout, A.B. 2007. Antimicrobial and antiviral effect of high-temperature short-time (HTST) pasteurization apllied to human milk. Breastfeed. Med. 2(1): 27–33. doi:10.1089/bfm.2006.0015.

Tian, P., Yang, D., Quigley, C., Chou, M., and Jiang, X. 2013. Inactivation of the tulane virus, a novel surrogate for the human norovirus. J. Food Prot. 76(4): 712–718. doi:10.4315/0362-028X.JFP-12-361.

Tomasula, P.M., Kozempel, M.F., Konstance, R.P., Gregg, D., Boettcher, S., Baxt, B., and Rodriguez, L.L. 2007. Thermal inactivation of foot-and-mouth disease virus in milk using high-temperature, short-time pasteurization. J. Dairy Sci. 90(7): 3202–3211. Elsevier. doi:10.3168/jds.2006-525.

Topping, J.R., Schnerr, H., Haines, J., Scott, M., Carter, M.J., Willcocks, M.M., Bellamy, K., Brown, D.W., Gray, J.J., Gallimore, C.I., and Knight, A.I. 2009. Temperature inactivation of Feline calicivirus vaccine strain FCV F-9 in comparison with human noroviruses using an RNA exposure assay and reverse transcribed quantitative real-time polymerase chain reaction-A novel method for predicting virus infectivity. J. Virol. Methods 156(1–2): 89–95. doi:10.1016/j.jviromet.2008.10.024.

Turner, C., Williams, S.M., and Cumby, T.R. 2000. The inactivation of foot and mouth disease, Aujeszky’s disease and classical swine fever viruses in pig slurry. J. Appl. Microbiol. 89(5): 760–767. doi:10.1046/j.1365-2672.2000.01174.x.

Underwood, M.A. 2013. Human Milk for the Premature Infant. Pediatr. Clin. North Am. 60(1): 189–207. doi:10.1016/j.pcl.2012.09.008.

Volk, M.L., Hanson, C. V, Israel-Ballard, K., and Chantry, C.J. 2010. Inactivation of Cell-Associated and Cell-Free HIV-1 by Flash-Heat Treatment of Breast Milk. JAIDS J. Acquir. Immune Defic. Syndr. 53(5): 665–666. doi:10.1097/QAI.0b013e3181ba47df.

Welch, J., Bienek, C., Gomperts, E., and Simmonds, P. 2006. Resistance of porcine circovirus and chicken anemia virus to virus inactivation procedures used for blood products. Transfusion 46(11): 1951–1958. doi:10.1111/j.1537-2995.2006.01003.x.

Welsh, J.K., Arsenakis, M., Coelen, R.J., and May, J.T. 1979. Effect of Antiviral Lipids, Heat, and Freezing on the Activity of Viruses in Human Milk. J. Infect. Dis. 140(3): 322–328. doi:10.1093/infdis/140.3.322.

Yue, C., Teitz, S., Miyabashi, T., Boller, K., Lewis-Ximenez, L., Baylis, S., and Blümel, J. 2019. Inactivation and Removal of Chikungunya Virus and Mayaro Virus from Plasma-derived Medicinal Products. Viruses 11(3): 234. doi:10.3390/v11030234.

Yunoki, M., Urayama, T., Yamamoto, I., Abe, S., and Ikuta, K. 2004. Heat sensitivity of a SARS-associated coronavirus introduced into plasma products. Vox Sang. 87(4): 302–303. doi:10.1111/j.1423-0410.2004.00577.x.

